# Identification of Blood-based Biomarkers for Early Stage Parkinson’s Disease

**DOI:** 10.1101/2020.10.22.20217893

**Authors:** Andrew Gao

## Abstract

Parkinson’s disease (PD) affects millions of people worldwide and causes symptoms such as bradykinesia and disrupted speech. Parkinson’s disease is known to be characterized by the mass death of dopaminergic neurons in the substantia nigra region. In the status quo, PD is often diagnosed at late stages because obvious motor symptoms appear after the disease has progressed far. It is advantageous to diagnose PD before the onset of motor symptoms because treatments are often more effective at early stages. While motor symptoms usually manifest when over 50% of dopaminergic neurons in the substantia nigra are already lost, molecular signatures of PD may be present at early stages in patient blood. This study aimed to analyze several gene expression studies’ data for commonly differentially expressed genes (DEGs) in the blood of early stage PD patients. 147 DEGs were identified in at least two out of three datasets and passed cut-off criteria. A protein interaction network for the DEGs was constructed and various tools were used to identify network characteristics and hub genes. PANTHER analysis revealed that the biological process “cellular response to glucagon stimulus” was overrepresented by almost 21 times among the DEGs and “lymphocyte differentiation” by 5.98 times. Protein catabolic processes and protein kinase functions were also overrepresented. ESR1, CD19, SMAD3, FOS, CXCR5, and PRKACA may be potential biomarkers and warrant further study. Overall, the findings of the present study provide insights on molecular mechanisms of PD and provide greater confidence on which genes are differentially expressed in PD. The results also are additional evidence for the role of the immune system in PD, a topic that is gaining interest in the PD research community.

## Introduction

The neurodegenerative disorder known as Parkinson’s disease (PD) affects approximately 6 million people worldwide [1]. PD is characterized by the loss of dopaminergic neurons in the substantia nigra region of the brain. Loss of these neurons causes symptoms ranging from tremors and affected posture to bradykinesia (slowed movements) [2]. Non-motion related symptoms, such as constipation and disruption of normal sense of smell, are also present in PD, often at early stages. Cognition and speech can also be affected, and PD complications include dementia and depression [3]. Currently, by the time most patients show obvious symptoms of PD, such as bradykinesia, PD has already progressed to an advanced stage with over half of dopaminergic neurons already lost [4]. In order to promote effective treatments, it is essential to diagnose PD before the advanced stage symptoms appear. However, this is relatively difficult as, as aforementioned, early stage symptoms of PD usually do not include outwardly apparent symptoms, but more subtle ones like constipation (which is not specific enough to PD to be especially useful). Some PD cases are due to genetics and could potentially be diagnosed via genetic screening, but the vast majority of PD cases arise spontaneously (idiopathic Parkinson’s disease) and genetic mutations may not be as apparent [5].

Thus, diagnostic tests and biomarkers for PD are in demand. There are a variety of potential avenues for biomarker discovery and researchers are actively exploring a diverse range of mediums, from saliva to cerebrospinal fluid [6–7]. Blood-based biomarkers are particularly attractive due to the relative non-invasive nature of the blood collection procedure, compared to, say cerebrospinal fluid extraction. Meanwhile, blood-based molecular biomarkers such as RNA confer more information about the disease state than an image.

Currently, there is still a lack of an accurate blood-based biomarker panel for early stage PD diagnosis. Besides applications for diagnosis tests, blood-based biomarkers could also be applied to developing novel treatments for Parkinson’s, perhaps someday allowing the development of therapeutics that can completely halt or even reverse PD progression.

Blood-based molecular biomarkers can be profiled using a variety of methods, such as RNA-seq and microarrays [8]. Both methods are used to perform gene expression studies. Microarray data has been used to identify biomarkers for other neurological disorders, such as Alzheimer’s disease [9]. Differential gene expression between PD patients and normal controls could provide detailed information about the mechanisms of PD, as well as of course offer valuable biomarkers for diagnosis. Previously, studies have identified PD biomarker genes through RNA-seq and microarray analysis. For example, one research group identified SRRM2 as a potential PD biomarker, using microarray data [10]. Another group established NAMPT as a potential PD blood biomarker [11].

While gene expression profiling methods are very useful, there are often substantial discrepancies between studies due to normal differences in methodology, sample characteristics, and more [12–13]. For example, the storage method and age of blood samples can have great effects on the contained RNA, many of which often degrade over time [14]. This means that genes identified as significant in one study may not be present in another. For greater confidence in biomarkers, it is important to identify biomarkers that routinely are significant in multiple studies. This decreases the likelihood that a result was due to an experimental flaw or coincidence.

In the present study, microarray data from three separate studies by different authors were analyzed to identify biomarkers for early stage PD. The studies all contained gene expression data based on the blood of patients who either had PD or were normal controls. Differentially expressed genes identified across studies were considered to be of special interest and their interactions were studied further.

## Methods

### Dataset Selection

The Gene Expression Omnibus (GEO) is an online NCBI repository containing public gene expression data from a variety of studies [15–16]. GEO was searched for datasets matching “blood”, “Parkinson’s”, “expression profiling by array”, and “*Homo sapiens”*. GSE6613, GSE54536, and GSE72267 were selected for this study [17–19]. In total, 94 PD patients and 45 normal controls were present across the three studies. All PD patients in the surveys were at early stages, according to the Hoehn and Yahr scale (ranges from 1 to 5). The average Hoehn and Yahr stage of the patients across the three studies is calculated to be 1.82 using the weighted average formula. Patients before stage 2 are considered to be in early stages of PD. All three studies were profiling gene expression from RNA extracted from patient blood. The studies used microarrays for data collection. Microarrays are a chip-based technology that uses oligonucleotide probes to capture strands of complementary DNA (cDNA) [20]. The cDNA is reverse transcribed from the mRNA present in the blood samples.

**Table 1:**
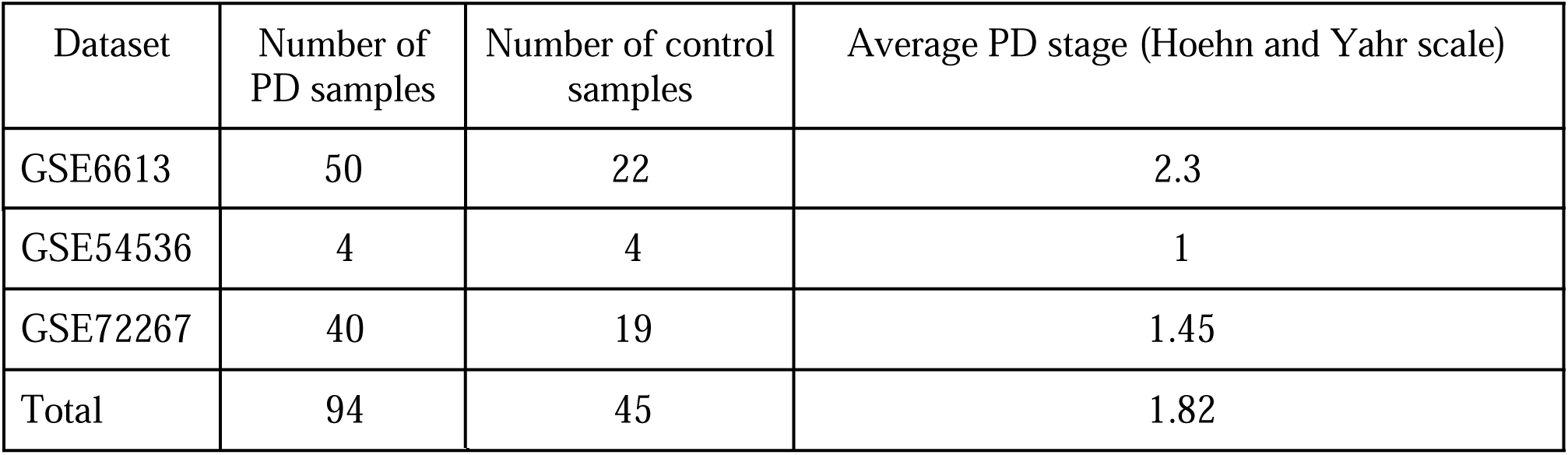
Dataset information for GSE6613, GSE54536, and GSE72267. There are 94 PD samples and 45 control samples total. The average PD stages of the patients is early: 1.82. At and after stage 3, PD becomes harder to treat and is phenotypically diagnosable via motor symptoms. Before stage 3, PD is still difficult to diagnose with conventional methods.

| Dataset | Number of PD samples | Number of control samples | Average PD stage (Hoehn and Yahr scale) |
| --- | --- | --- | --- |
| GSE6613 | 50 | 22 | 2.3 |
| GSE54536 | 4 | 4 | 1 |
| GSE72267 | 40 | 19 | 1.45 |
| Total | 94 | 45 | 1.82 |

### Differential Expression Analysis

The three datasets were analyzed using the tool GEO2R which is provided by the Gene Expression Omnibus [15]. GEO2R is a browser-based software that processes gene expression values and outputs a table of differentially expressed genes (DEGs) between two user-defined groups (PD and control, in this case). In total, several thousand genes were deemed statistically significant by GEO2R for each dataset. T tests were used to determine p values. GEO2R was also used to calculate fold changes for each gene in the context of PD expression values vs. control values. The fold change is a ratio of the average expression value of a gene in one group divided by the average expression value in a different group. Fold change can be greater or less than one; some genes are overexpressed while others are underexpressed in PD compared to normal controls.

GEO2R was also used to verify a normal distribution of gene expression values. No outliers were present.

After the lists of DEGs were obtained, they were processed in Excel. DEGs with p values of greater than 0.05 were removed and the remaining DEGs were sorted into two categories: overexpression and underexpression. Some genes were less expressed in PD than controls while others were more highly expressed in PD. Next, only genes with fold change of greater than 1.25 were kept.

### Identification of Common Genes

The online tool Jvenn was used to create a Venn diagram of the genes contained in each of the three studies [21]. Genes present in at least two out of three studies were saved while genes found in only one were removed from the list.

### Network Analysis

The final list of DEGs was analyzed in STRING to construct a network of the protein-protein interactions of the DEG products. STRING, the Search Tool for the Retrieval of Interacting Proteins, scans numerous databases to create a graph of input genes and how their protein products interact [22]. STRING also calculates statistically significant Gene Ontology processes and pathways. A tab separated value file containing the gene graph in text form was exported and imported into Cytoscape [23]. Cytoscape is a desktop-based tool that can perform more powerful analyses and functions on networks than STRING. The Network Analyzer tool in Cytoscape was used to calculate metrics of the gene network such as node degree and clustering coefficient. Genes with a degree of >= 8 were considered hub genes. The degree is the number of genes connected to a certain gene.

### Gene Ontology Analysis

The Gene Ontology (GO) is a database that contains information on biological pathways, processes, components, and the genes that affect them [24–25]. GO has a built in PANTHER tool that accepts a list of genes and identifies biological entities that the genes are relevant in [26]. The list of DEGs was submitted to PANTHER and enriched biological processes and molecular functions were identified.

## Results

GSE6613, GSE54536, and GSE72267 were analyzed in GEO2R and the lists of DEGs were downloaded. Gene expression value distribution was confirmed to be normal, with no outliers. Cut-off criteria for genes were p value <0.05 and fold change > 1.25.

In GSE6613, 828 genes passed the criteria, in GSE54536, 2370, and in GSE72267, 259.

There was substantial overlap between studies. 145 genes were found in two out of three datasets and two genes were found in all three. Those two genes were TMEM19 and PPP2R3A.

TMEM19 codes for a transmembrane protein involved in protein binding, but not much else is known [27]. PPP2R3A codes for an enzyme, protein phosphatase 2, that is shown to be involved in negatively regulating cell growth and division [28–29].

The 147 genes that passed all cut-off criteria and were found in a minimum of two datasets were submitted to the STRING tool. 146 out of 147 genes were successfully mapped and the following graph was created. Solitary genes were hidden to improve clarity. The minimum confidence score for whether a protein interaction existed was set to 0.400. There were 134 connections between proteins instead of an expected 105, indicating that the network has significantly more interactions than predicted of a random sample (p value = 0.004).

STRING identified several biological processes as significant among the list of genes.

Next, Cytoscape software was used to analyze network characteristics, namely node degree. 6 genes were considered hub genes because they had at least a node degree of 8.

**Table 2:**
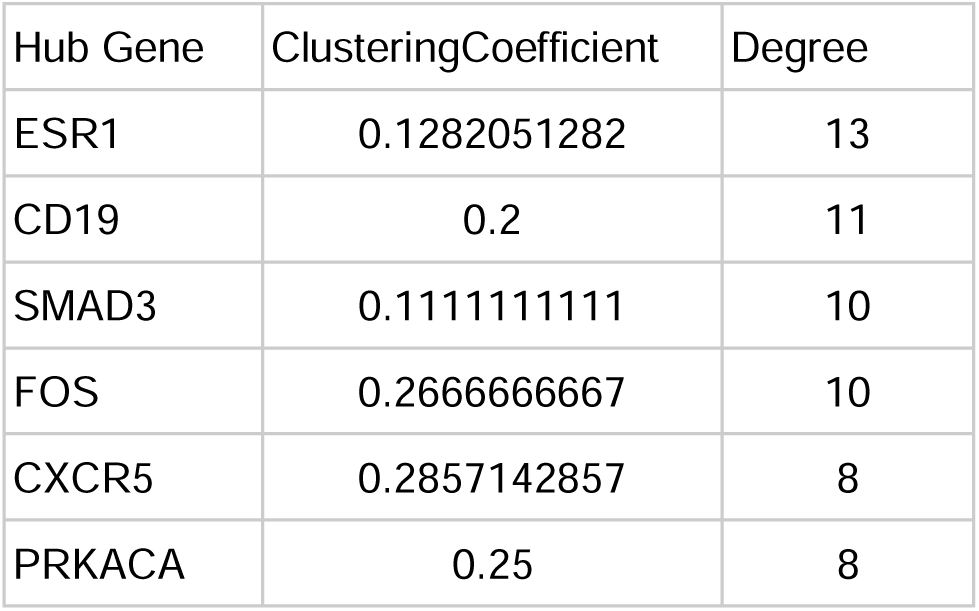
Cytoscape calculated clustering coefficients and node degrees for the genes in the network. The genes with node degree of at least 8 are presented: ESR1, CD19, SMAD3, FOS, CXCR5, and PRKACA. The local clustering coefficient is a metric that measures how clustered nodes are around a single node, in this case, the hub genes. A clustering coefficient of 1 indicates the maximum possible density of a cluster around a node while 0 is the least dense.

13 gene products were found by STRING to interact with ESR1. ESR1 codes for an estrogen receptor that plays a role in hormonal signaling, DNA binding, and transcription. ESR1 is disrupted in diseases such as breast cancer and osteoporosis [30]. ESR1 was downregulated in GSE72267 and GSE54536, but upregulated in GSE6613.

Finally, Gene Ontology (GO)’s PANTHER tool was used to analyze the gene list. PANTHER was set to:

1. Annotation Data Set: GO biological process complete
2. Test Type: Fisher’s Exact
3. Correction: Calculate False Discovery Rate (FDR)

PANTHER identified a number of statistically significant biological processes. Most notably, the process “cellular response to glucagon stimulus” was enriched by 20.98 times, with p value = 6.23E-05 and false discovery rate of 0.00495. Enrichment is the number of genes in the list that belong to a specific biological process divided by the expected number of genes that would belong if taken from a random sample of genes. A higher enrichment value indicates that a greater amount of genes in a list are involved in a certain GO process.

“Regulation of proteolysis involved in cellular protein catabolic process”, “lymphocyte differentiation”, “regulation of immune system process”, and “cellular protein modification process” were also enriched.

**Table 3:** PANTHER outputs a table showing the various biological processes that are over or under-represented among the list of genes. The table is ranked by largest fold enrichment. All presented processes are statistically significant. GO accession terms are provided for each process. The table displays the number of genes in the list that are found in a process, the expected number of genes, the enrichment, the p value, and the false discovery rate (FDR).

| GO biological process complete | Genes in List | Expected Genes | Fold Enrichment | p value | FDR |
| --- | --- | --- | --- | --- | --- |
| cellular response to glucagon stimulus (GO:0071377) | 4 | 0.19 | 20.98 | 6.23E-05 | 4.95E-02 |
| regulation of proteolysis involved in cellular protein catabolic process (GO:1903050) | 10 | 1.55 | 6.44 | 5.22E-06 | 1.66E-02 |
| lymphocyte differentiation (GO:0030098) | 10 | 1.69 | 5.92 | 1.06E-05 | 2.41E-02 |
| regulation of cellular protein catabolic process (GO:1903362) | 10 | 1.78 | 5.6 | 1.67E-05 | 2.66E-02 |
| regulation of protein catabolic process (GO:0042176) | 14 | 2.76 | 5.08 | 1.01E-06 | 1.61E-02 |
| leukocyte differentiation (GO:0002521) | 11 | 2.41 | 4.56 | 3.95E-05 | 3.93E-02 |
| lymphocyte activation (GO:0046649) | 12 | 2.68 | 4.47 | 2.11E-05 | 2.80E-02 |
| immune system development (GO:0002520) | 16 | 4.6 | 3.48 | 1.84E-05 | 2.67E-02 |
| hemopoiesis (GO:0030097) | 14 | 4.03 | 3.47 | 6.47E-05 | 4.90E-02 |
| hematopoietic or lymphoid organ development (GO:0048534) | 15 | 4.36 | 3.44 | 3.87E-05 | 4.11E-02 |
| regulation of immune system process (GO:0002682) | 27 | 11.86 | 2.28 | 5.25E-05 | 4.64E-02 |
| cellular protein modification process (GO:0006464) | 44 | 21.75 | 2.02 | 3.21E-06 | 2.56E-02 |
| protein modification process (GO:0036211) | 44 | 21.75 | 2.02 | 3.21E-06 | 1.70E-02 |

Many GO molecular functions were altered as well. For example, the cAMP-dependent protein kinase activity function was enriched by 65.26 times which is quite a large enrichment. Protein kinase functions appear to be commonly disrupted in PD.

**Table 4:** PANTHER outputs a table showing the various molecular functions that are over or under-represented among the list of genes. All presented functions are statistically significant. GO accession terms are provided for each function. The table displays the number of genes in the list that are found in a function, the expected number of genes, the enrichment, the p value, and the false discovery rate (FDR).

| GO molecular function complete | Genes in list | Expected Genes | Fold Enrichment | p value | FDR |
| --- | --- | --- | --- | --- | --- |
| cAMP-dependent protein kinase activity (GO:0004691) | 4 | 0.06 | 65.26 | 1.37E-06 | 6.51E-03 |
| cyclic nucleotide-dependent protein kinase activity (GO:0004690) | 4 | 0.07 | 53.4 | 2.58E-06 | 6.15E-03 |
| AMP-activated protein kinase activity (GO:0004679) | 3 | 0.07 | 44.05 | 8.25E-05 | 3.92E-02 |
| protein kinase A regulatory subunit binding (GO:0034237) | 4 | 0.18 | 21.75 | 5.48E-05 | 3.26E-02 |
| SMAD binding (GO:0046332) | 6 | 0.54 | 11.01 | 2.60E-05 | 2.06E-02 |
| beta-catenin binding (GO:0008013) | 6 | 0.59 | 10.13 | 4.05E-05 | 2.75E-02 |
| ubiquitin-like protein ligase binding (GO:0044389) | 12 | 2.18 | 5.51 | 2.78E-06 | 4.41E-03 |
| ubiquitin protein ligase binding (GO:0031625) | 11 | 2.05 | 5.37 | 9.29E-06 | 1.10E-02 |
| transcription coactivator activity (GO:0003713) | 9 | 1.77 | 5.08 | 9.23E-05 | 3.99E-02 |
| catalytic activity, acting on a protein (GO:0140096) | 34 | 15.49 | 2.2 | 9.92E-06 | 9.44E-03 |
| enzyme binding (GO:0019899) | 32 | 15.7 | 2.04 | 8.22E-05 | 4.35E-02 |

## Discussion

Parkinson’s disease is a prevalent neurodegenerative disease that is difficult to diagnose at early stages due to the lack of many visible motor symptoms. A blood-based gene biomarker approach may be more fruitful in diagnosing patients earlier so they may receive more effective treatment.

Analysis of three microarray datasets on PD gene expression yielded 147 differentially expressed genes that are found across studies. These genes pass criteria for p value and fold change and are attractive for further research as they are both biologically relevant and unlikely to be irreproducible. While 145 genes were present in two of three studies, only two, TMEM19 and PPPR23A, were found in all datasets, demonstrating how variable gene expression studies can be (and highlighting the need for this study).

ESR1, CD19, SMAD3, FOS, CXCR5, PRKACA all had at least eight interactions with other genes, indicating that they may play an important role in PD. ESR1, SMAD3, PRKACA, and FOS were located in the center of a cluster of other genes, such as ITCH and NOTCH2.

ESR1 was shared across 2 studies and was a hub gene with degree 13. Variants of ESR1 have been linked to different levels of susceptibility to Parkinson’s. Estrogen protects the nigrostriatal pathway, which is perturbed by PD. This may partially explain why men are more likely to develop PD than women, as another study found [30].

CD19, a gene with 11 protein interactions out of the 146 other genes, codes for a transmembrane protein. CD19 is highly expressed in B cells and especially during B cell development up to the point of the end of differentiation. CD19 is essential for maintaining the function of B cell receptor transmembrane proteins that regulate how the B cell grows or dies [31]. CD19 is interesting as its function is involved with the Gene Ontology process of “leukocyte differentiation” that was identified in the PANTHER analysis.

SMAD3 codes for a protein that transmits signals to the nucleus from the cell surface, specifically for the transforming growth factor-beta signaling pathway [32]. SMAD3 and other SMAD family proteins merge to form a complex that can control gene expression by attaching to DNA. SMAD proteins affect cell division, apoptosis, and movement, amongst other things. The SMAD3 protein directly interacts with ESR1 and FOS, as shown in Figure 4.

**Figure 1:**
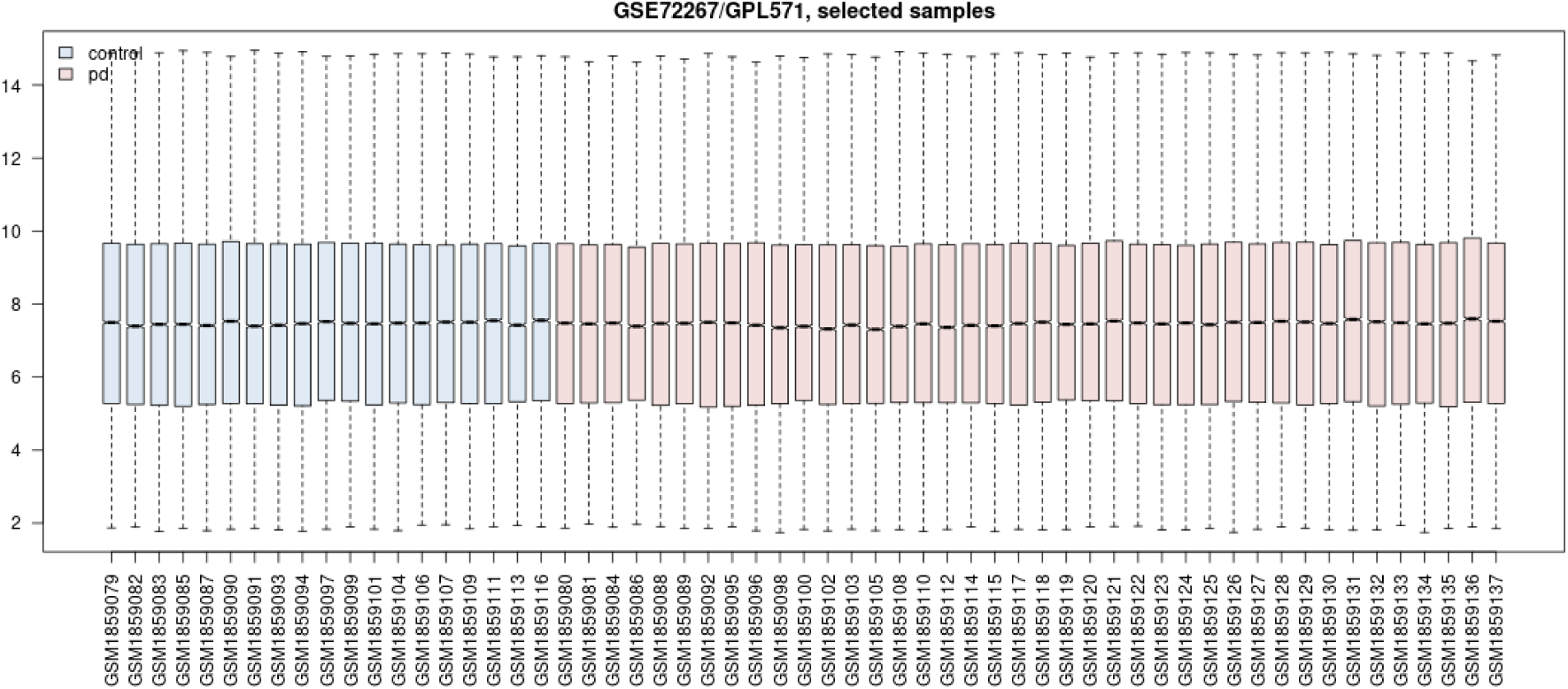
Gene expression value distribution for GSE72267. There are no units for the y axis. Each box plot represents the gene expression values of one patient sample.

**Figure 2:**
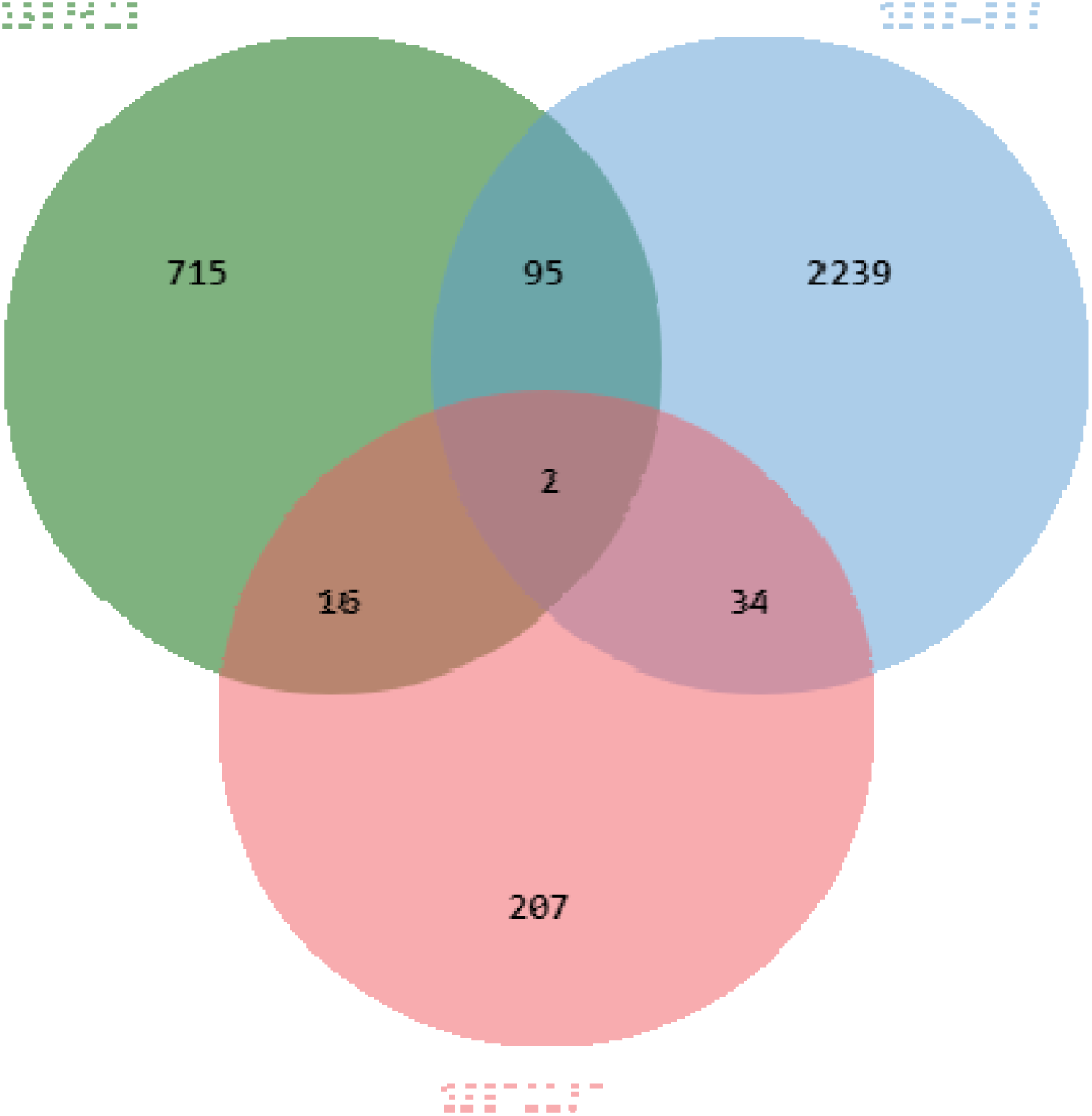
The Venn diagram shows the number of genes that are shared between GSE6613, GSE54536, and GSE722667. For example, 95 genes were differentially expressed in both GSE54536 and GSE6613.

**Figure 3:**
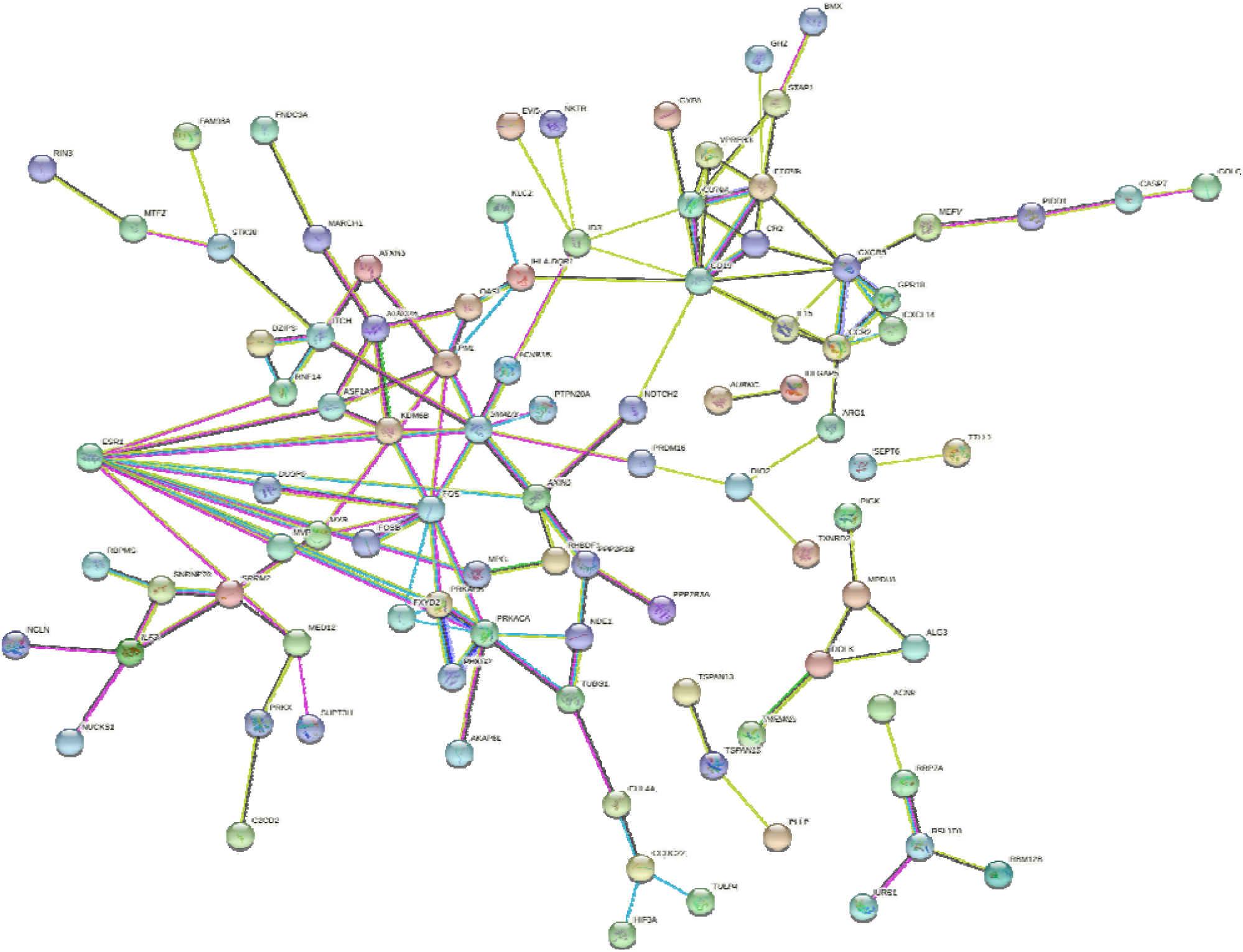
STRING network of protein interactions between 146 differentially expressed genes. ESR1, a blue node in the left of the map, is shown to have many interactions with other proteins. The colors of the nodes are arbitrary. The colors of the edges between nodes indicate how the interaction was determined. A pink line, for example, means that the interaction was experimentally demonstrated. Isolated genes are removed from the diagram for clarity.

**Figure 4:**
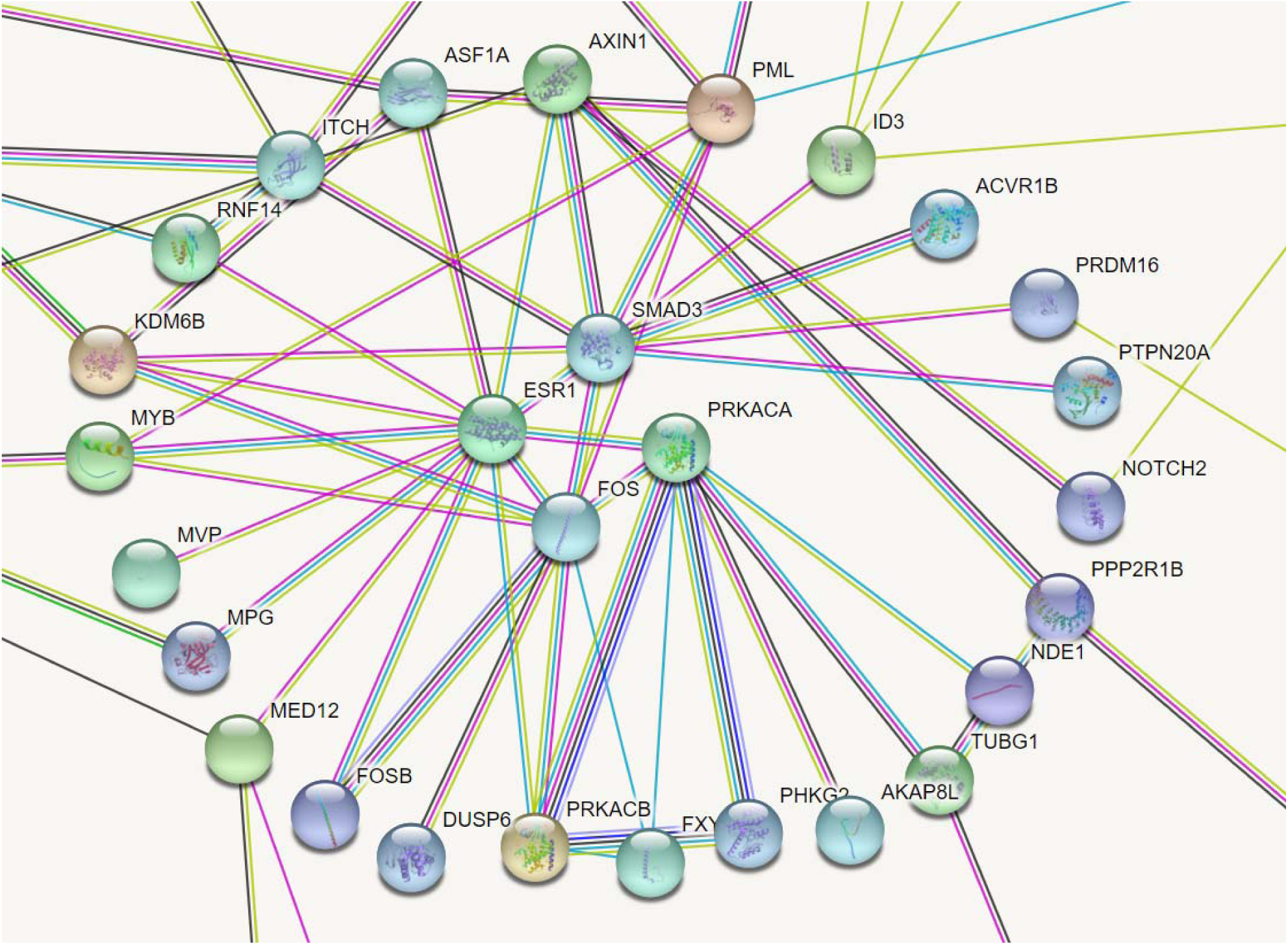
STRING network focused on SMAD3, ESR1, FOS, and PRKACA, four hub genes. There are many protein intersections between these four genes and surrounding genes.

**Figure 5:**
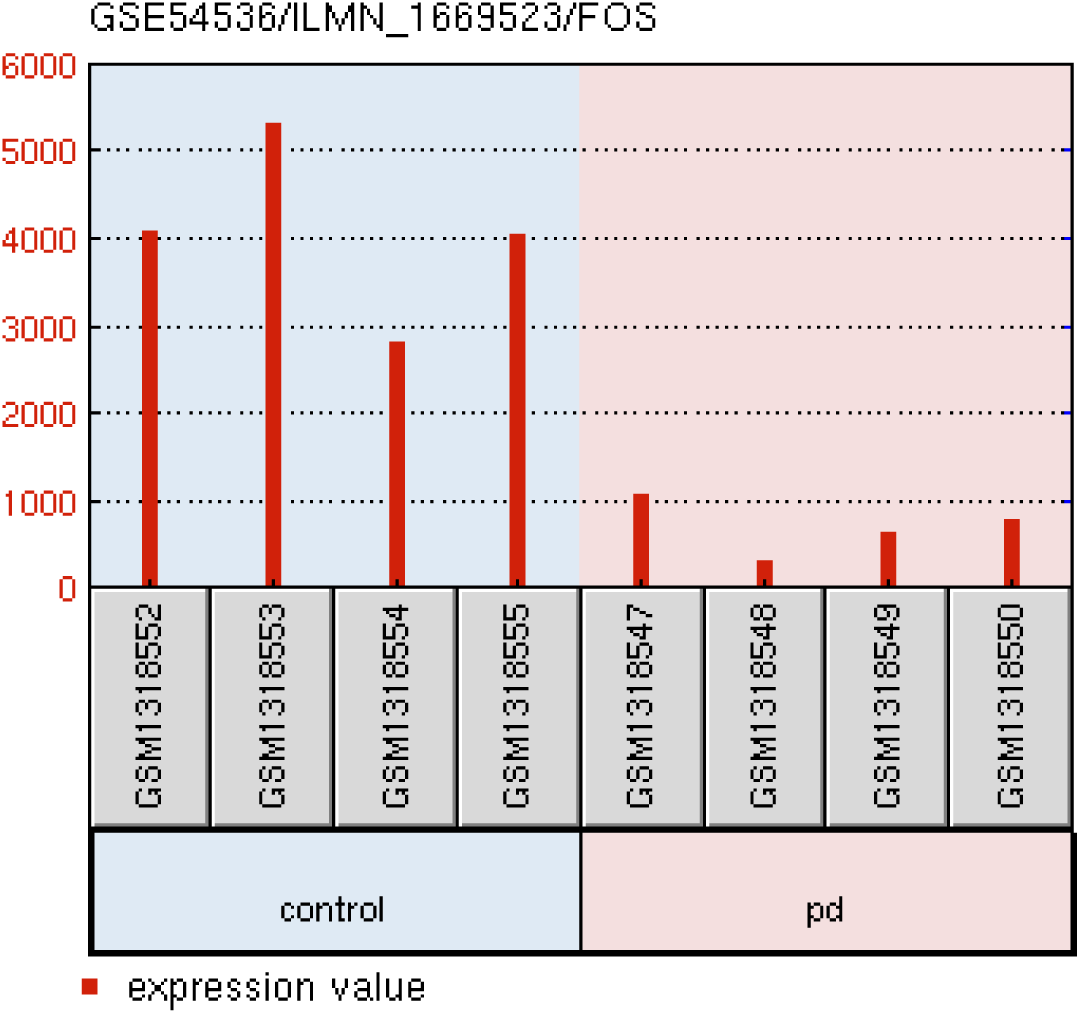
FOS expression values for GSE54536 samples. The average fold change is .169 with p = 0.00023. FOS is underexpressed in PD patients.

**Figure 6:**
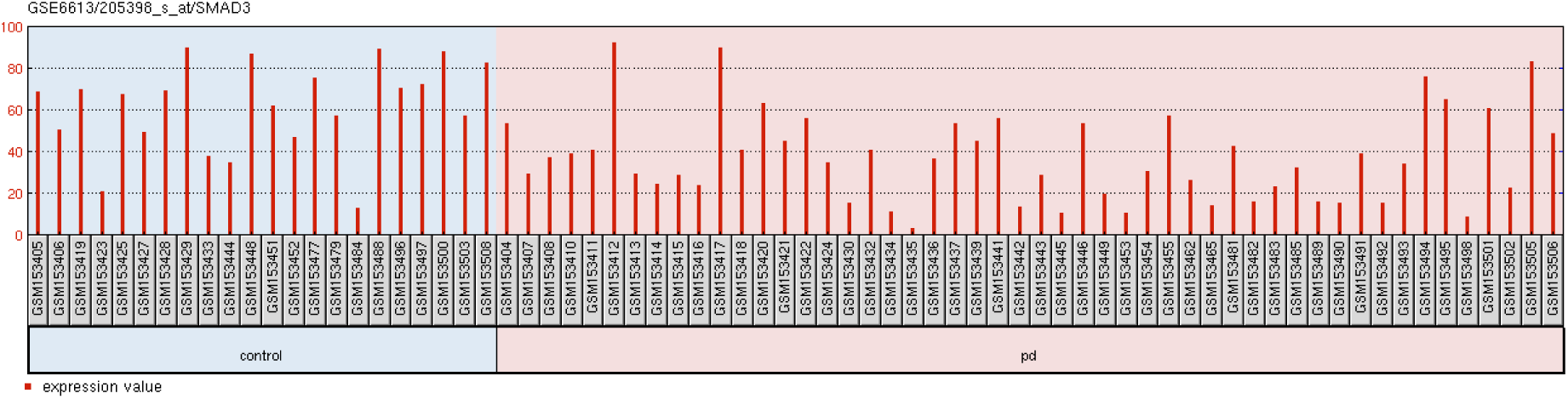
SMAD3 expression values for GSE6613 samples. The average fold change is .549 with p = 0.00021.

While the hub genes are promising, other genes among the 147 may be of interest as well. The complete list is attached at the end of this report.

“Cellular response to glucagon stimulus” genes were extremely over-represented in the list of DEGs (20.98 times enriched). The Glucagon-like Peptide-1 Receptor is currently being explored for new strategies for treating PD. Stimulating the Glucagon-like Peptide-1 Receptor has neuroprotective effects [33]. Future research could be done on harnessing the effects of such stimulation for PD therapeutics.

Genes involved in “lymphocyte differentiation” were also over-represented by 5.92 times. This implies that lymphocytes are somehow dysregulated in PD, drawing attention to the fact that PD may be associated with immune system dysfunction. To date, not much literature has described the role of lymphocytes in PD so the findings of this study may shed light on the intersection between immunology and PD. While there is relatively little experimental information, in 2018, Kozina et al. found that mice with LRRK2 mutations experienced neurodegeneration when immune cell factors from the peripheral blood crossed the blood brain barrier and caused neuroinflammation [34]. LRRK2 is a famous mutation implicated in PD.

Numerous unique molecular functions concerning the function of protein kinases were found amongst the list of genes and were often significantly over-represented. In the case of “cAMP-dependent protein kinase activity”, four genes in the list were part of the Gene Ontology function despite the rarity of genes involved in the function. The function was over-represented by 65.26 times (p = 1.37E-06). “cyclic nucleotide-dependent protein kinase activity”, “AMP-activated protein kinase activity”, and “protein kinase A regulatory subunit binding” were also over-represented. The importance of protein kinases in PD suggests that avenues for treatment may include designing protein kinase inhibitors. Further research should be conducted on the implications of protein kinase dysregulation in PD and how it can be manipulated.

Strengths of this study include the rigor applied to differentially expressed gene selection. To qualify for the final list, a gene had to have a fold change of at least 1.25, a p value of < 0.05, and be present in ⅔ datasets. Another strength is the sample size. PD can vary considerably at the molecular level between patients. By considering nearly one hundred PD patients, there is greater assurance of obtaining a general view of the gene expression of PD than with just 4 samples, for example, in the case of GSE54536. Additionally, another strength of this study is that the samples analyzed are all from early stage PD (Hoehn-Yahr average 1.82) compared to other studies where samples were often in late stages at the time of collection. At late stages, there are often different gene expression differences that may not necessarily be present at early stages.

Limitations of this study are that no normalization was performed between datasets (although this is common with cross-study gene expression analyses). Additionally, there was no distinction between male and female samples in the analysis. There may be significant differences in gene expression and PD pathology between sexes that were ignored in this study. Biomarkers that may work well for females may not be effective for males. Another important caveat to note is that the research only compared healthy normal controls and early stage PD patients. However, PD patients were not compared with other neurological controls. Thus, it is not known whether the DEGs found are specific to PD. GSE6613 contains gene expression data from non-PD neurological disorder controls, so it follows that the next research step is to evaluate the biomarkers identified in this study and verify if they are differentially expressed in PD vs other neurological disorders. Finally, the blood brain barrier can restrict many molecules from crossing from the brain to the bloodstream so our analysis is constrained to downstream effects rather than observing direct changes in dopaminergic neurons, which are out of reach in the brain [35]. The results of this study should be confirmed in vitro if possible.

The list of DEGs identified may be useful as diagnostic markers for non-invasive PD tests and they may also be useful targets for novel treatments for PD. The present study elucidates several biological processes, such as the cellular response to glucagon stimulus and lymphocyte differentiation, that could be avenues for research in alleviating PD symptoms. Currently, little is known about the role of the immune system in PD development, however, there is growing interest. Dzamko et al. describes that changes to the peripheral immune system can affect the onset of PD [36]. This study’s finding that lymphocyte differentiation genes are over-represented in PD patient blood provides further evidence for Dzamko’s findings.

Future research directions include finding genes that can distinguish between PD and other neurodegenerative conditions and exploring epigenetic factors such as miRNAs and histone modifications and their interactions with genes. Examining the role of glucagon stimulus and lymphocyte differentiation in PD may also yield important information.

## Supporting information

Supplemental List of Genes

## Data Availability

The data is publicly available from the respective Gene Expression Omnibus studies from which they were retrieved. I do not claim credit for the data. Other researchers obtained that data and credit goes to them (study accession pages are linked).
Supplemental
STRING network:
https://version-11-0b.string-db.org/cgi/network?networkId=bns9q9ZNmuqR
List of 147 Differentially Expressed Genes
https://docs.google.com/spreadsheets/d/1kAj7B2oXeNSK-Bha7Xo14IRv_Z1xTGCcdwOxRYdWem8/edit?usp=sharing
Venn Diagram Results
https://docs.google.com/spreadsheets/d/1g4-k2lGj78hG1rLhMsQbK77MQ-TBe9QQNAADhLfFDlI/edit?usp=sharing

https://www.ncbi.nlm.nih.gov/geo/query/acc.cgi?acc=GSE6613

https://www.ncbi.nlm.nih.gov/geo/query/acc.cgi?acc=gse54536

https://www.ncbi.nlm.nih.gov/geo/query/acc.cgi?acc=gse72267

## Acknowledgements

Thank you to my mentor, Wendy Slijk, for general advice.

## Supplemental

### STRING network

https://version-11-0b.string-db.org/cgi/network?networkId=bns9q9ZNmuqR

### List of 147 Differentially Expressed Genes

https://docs.google.com/spreadsheets/d/1kAj7B2oXeNSK-Bha7Xo14IRv_Z1xTGCcdwOxRYdWem8/edit?usp=sharing

### Venn Diagram Results

https://docs.google.com/spreadsheets/d/1g4-k2lGj78hG1rLhMsQbK77MQ-TBe9QQNAADhLfFDlI/edit?usp=sharing

